# Pre-admission polypharmacy burden and intensive care unit outcomes in patients with sepsis: A retrospective cohort study using the MIMIC-IV-ED linked database

**DOI:** 10.64898/2026.05.12.26352808

**Authors:** Farhana Haque, Mahmudul Hasan

## Abstract

**Purpose:** Polypharmacy is highly prevalent among critically ill patients, yet it’s independent impact on intensive care unit (ICU) outcomes in sepsis remains critically unexplored. We aimed to evaluate whether pre-admission polypharmacy independently predicts ICU mortality and provides incremental prognostic value using the medication reconciliation module of the MIMIC-IV-ED linked database. Materials and

**Methods:** We conducted a retrospective cohort study of 3,347 adults admitted to the ICU who met Sepsis-3 criteria. Pre-admission polypharmacy was categorized as none (0-4), standard (5-9), or high (≥10 medications). Multivariable logistic regression, propensity score matching, and reclassification analyses (NRI/IDI) were performed. The primary outcome was in-hospital ICU mortality.

**Results:** High polypharmacy was present in 58.9% of patients. Crude ICU mortality increased sequentially: 18.5% (none), 26.0% (standard), and 27.5% (high; p < 0.001). After multivariable adjustment, high polypharmacy independently predicted in-hospital ICU mortality (aOR 1.45, 95% CI (1.10-1.91)), and 28-day mortality (aOR 1.47). Drug-class analysis identified statins as significantly protective (aOR 0.56), whereas RAS blockers combined with diuretics increased acute kidney injury risk (aOR 1.49). Propensity matching confirmed the primary mortality association (matched aOR 1.28).

**Conclusions:** By utilizing the ED medication reconciliation table, this study proves high polypharmacy represents a distinct ‘pharmacologic frailty’, independent of acute severity. Available instantly at triage, this zero-latency metric provides significant early prognostic value (SOFA NRI = 0.24) and identifies actionable high-risk interactions (e.g., RAS blockers plus diuretics) for immediate, targeted pharmacist-led intervention upon ICU admission.

## 1. Introduction

Sepsis, defined as life-threatening organ dysfunction caused by a dysregulated host response to infection, remains the leading cause of ICU mortality worldwide, responsible for approximately 11 million deaths annually and accounting for nearly one-fifth of all global deaths [1]. While in-hospital interventions such as timely antibiotic administration, evidence-based fluid resuscitation, and source control have received extensive investigation, the role of pre-admission patient characteristics in shaping critical illness trajectories is comparatively under-studied. Among pre-admission factors, medication burden represents a clinically accessible yet systematically neglected exposure.

Polypharmacy, most commonly defined as the concurrent use of five or more medications, affects 35–60% of adults presenting to emergency departments in developed countries, and is especially prevalent among older adults with multiple chronic conditions [2, 3]. High polypharmacy (≥10 medications) is associated with drug-drug interactions, immune modulation, altered pharmacokinetics, and frailty [4]. In non-critical settings, polypharmacy predicts adverse outcomes including hospitalisation, falls, and death [5]. The biological plausibility for polypharmacy worsening sepsis outcomes is compelling: renin-angiotensin system (RAS) blockers and diuretics may impair renal autoregulation under sepsis-induced hypoperfusion; anticoagulants alter the coagulation-inflammation interface; corticosteroids suppress innate immunity; and the aggregate medication burden itself may reflect a phenotype of multimorbidity-related physiologic fragility not fully captured by standard severity scores [6]. Despite this biological rationale, no published study has leveraged the MIMIC-IV-ED medication reconciliation table (*medrecon*), which prospectively documents all home medications at the point of ED triage, to examine whether pre-admission polypharmacy independently predicts ICU mortality in septic patients.

We therefore conducted a large retrospective cohort study using linked MIMIC-IV-ED and MIMIC-IV data to test the hypothesis that pre-admission high polypharmacy (≥10 medications) is independently associated with increased ICU mortality in sepsis, after adjustment for established severity predictors. We additionally examined the doseresponse relationship, drug-class contributions, and incremental predictive value of polypharmacy beyond existing severity scores.

## 2. Methods

### 2.1 Study design and data source

We conducted a retrospective cohort study using MIMIC-IV-ED (v2.2) linked to MIMIC-IV (v2.2), two freely accessible de-identified electronic health record databases derived from Beth Israel Deaconess Medical Center (BIDMC), Boston, Massachusetts [7, 8]. MIMIC-IV-ED contains a *medrecon* table documenting all medications patients were taking prior to their ED presentation, recorded by nursing staff during standardised triage intake. Databases are linked via a common patient identifier (*subject_id*), permitting ED medication records to be joined to subsequent ICU admission data, laboratory values, and mortality outcomes. The study was conducted in accordance with the PhysioNet data use agreement; institutional review board approval was not required as all data are de-identified per HIPAA Safe Harbor.

### 2.2 Study population and cohort construction

Starting from 425,087 total ED stays in MIMIC-IV-ED, we identified 203,016 with a linked hospital admission and 35,307 with a subsequent ICU stay. After retaining the earliest ICU stay per hospital admission, 31,867 unique patient-admissions remained. We then applied sequential exclusions: Sepsis-3 criteria (ICD-10 A40.x/A41.x or ICD-9 038.x/995.91/995.92) excluded 25,502 non-sepsis patients (n = 6,365 remaining); unavailable medication reconciliation data excluded 1,683 (n = 4,682); restriction to first ICU admission per patient excluded 607 repeat admissions (n = 4,075); ICU stay <24 hours excluded 716 (n = 3,359); and death within 24 hours of ICU admission excluded 12, yielding a final analytic cohort of 3,347 patients (Fig. 1).

**Figure 1:**
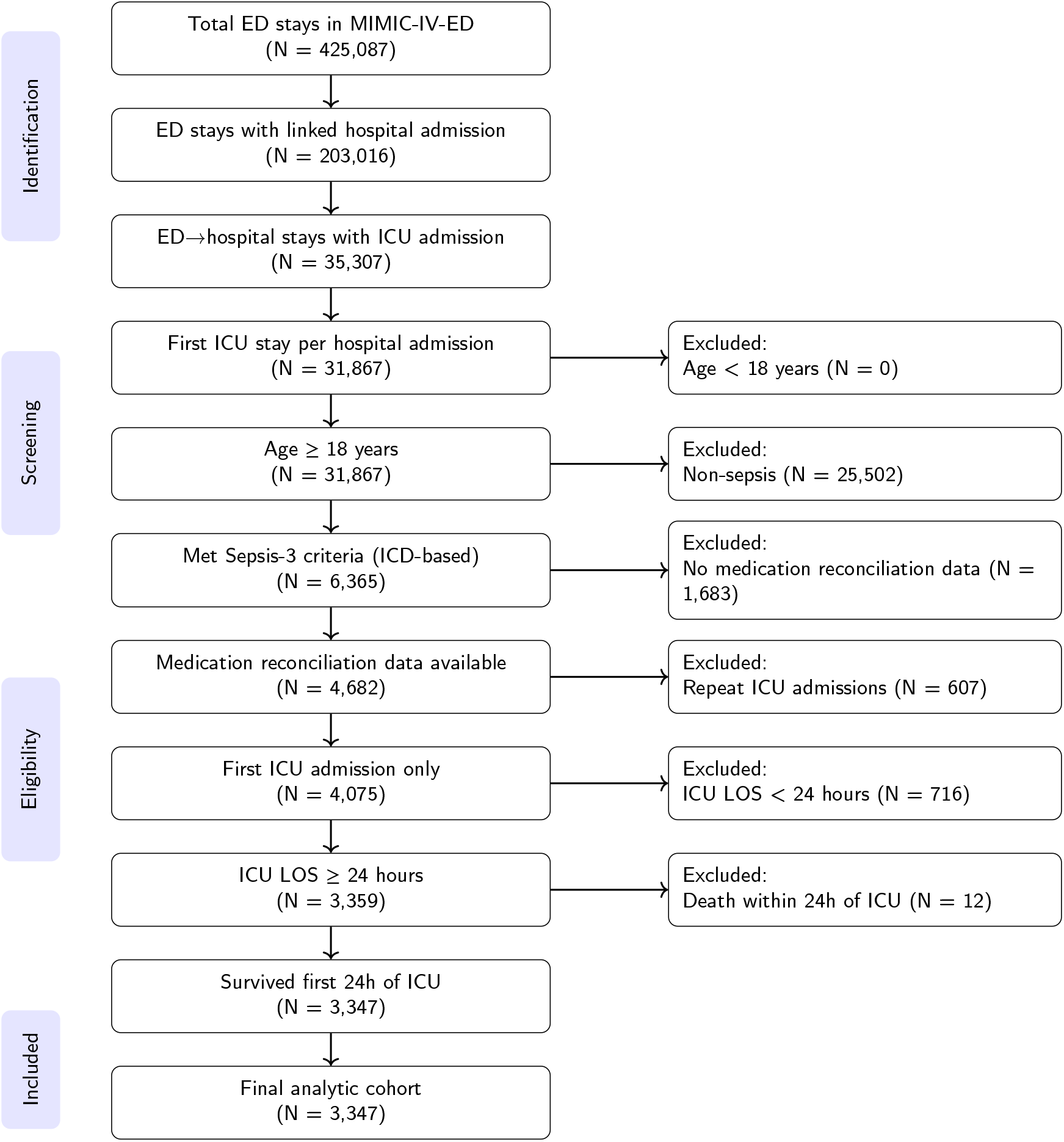
Study flow diagram. Sequential application of inclusion and exclusion criteria from 425,087 total ED stays in MIMIC-IV-ED to the final analytic cohort of 3,347 patients. Key exclusion steps: 25,502 non-sepsis by ICD criteria; 1,683 without medication reconciliation data; 607 non-first admissions; 716 with ICU LOS <24 hours; 12 deaths within 24 hours of ICU admission.

### 2.3 Primary exposure

The primary exposure was the total count of distinct medications documented in the *medrecon* table per patient, deduplicated by drug name using the *etcdescription* field. Polypharmacy was classified as: none (0–4 medications), standard polypharmacy (5–9 medications), and high polypharmacy (≥10 medications), following established clinical thresholds [2, 9]. Medication count was also analysed as a continuous variable in spline regression. For drug-class subanalyses, home medications were grouped into nine categories using the *etccode* ontology field: RAS agents (ACE inhibitors, ARBs), beta-blockers, statins, anticoagulants, systemic corticosteroids, immunosuppressants, diuretics, antidiabetic agents, and opioids/antipsychotics.

### 2.4 Outcomes

The primary outcome was in-hospital all-cause ICU mortality, defined as death during the index hospitalisation, determined by linking the *patients*.*dod* field to hospital admission and discharge timestamps. Secondary outcomes were: (1) 28-day all-cause mortality; (2) ICU length of stay (LOS; hours from *icustays*.*intime* to *icustays*.*outtime*); (3) hospital LOS (days from *admissions*.*admittime* to *admissions*.*dischtime*); and (4) new-onset AKI per KDIGO criteria, defined as a serum creatinine rise ≥0.3 mg/dL within 48 hours or ≥1.5× baseline within 7 days of ICU admission, derived from the *labevents* table [10].

### 2.5 Covariates

Covariates extracted *a priori* included: age, sex, race/ethnicity, insurance type, SOFA score (computed from chartevents and labevents within the first 24 hours of ICU admission), Charlson Comorbidity Index (CCI; from ICD-10 codes [11]), first serum lactate within 6 hours of ICU admission (dichotomised at >2 mmol/L), septic shock (vasopressor requirement plus lactate >2 mmol/L [1]), mechanical ventilation within 24 hours, primary infection source (pulmonary, urinary, abdominal, bloodstream, other derived from ICD codes), and time from ED presentation to ICU admission (hours).

### 2.6 Statistical analysis

Baseline characteristics were compared across polypharmacy groups using the Kruskal-Wallis test (continuous variables) and chi-square test (categorical variables). Multivariable logistic regression with all prespecified covariates simultaneously entered was used for the primary outcome. Restricted cubic spline (RCS) regression with four knots at the 5th, 35th, 65th, and 95th percentiles of medication count assessed dose-response, with non-linearity tested by likelihood ratio test [12]. Propensity scores for high polypharmacy were estimated by logistic regression incorporating all covariates, followed by 1:1 nearest-neighbour propensity score matching with replacement (caliper = 0.2 SD of the logit of the propensity score); matching with replacement was used given that the control pool (n = 1,377) was smaller than the treated group (n = 1,970), which precludes matching without replacement. Covariate balance was assessed by standardised mean differences (SMD < 0.10). Incremental predictive value was quantified by continuous net reclassification improvement (NRI) and integrated discrimination improvement (IDI) for polypharmacy added to SOFA-only and SOFA+CCI models. A drug class sub-analysis examined individual medication classes within the high polypharmacy stratum. Missing data for variables with <5% missingness were handled by complete-case analysis; variables with 5–20% missing data would have been imputed using multiple imputation by chained equations [13]. If a subgroup model failed to converge due to complete or near-complete separation, the subgroup would be reported as non-estimable with the reason documented. All analyses used Python 3.13 (pandas, statsmodels, scikit-learn) and R 4.3. A two-sided p < 0.05 was considered significant.

## 3 Results

### 3.1. Study population and cohort flow

The final analytic cohort comprised 3,347 patients (Fig. 1). The median age was 65 years; 54.3% were male; and 70.9% identified as White. Mechanical ventilation within 24 hours was nearly universal (99.0%), reflecting the severity of the ICU-admitted sepsis population. Septic shock was present in 34.0%. The overall in-hospital ICU mortality was 25.9% (867/3,347) and 28-day mortality was 30.7% (1,029/3,347). Median ICU LOS was 2.8 days (IQR 1.8–5.3) and median hospital LOS was 9.8 days (IQR 5.8–17.6). New-onset AKI by KDIGO criteria occurred in 50.6% of patients. Lactate >2 mmol/L was present in 70.2% of patients overall (72.2% none, 71.8% standard, 68.9% high polypharmacy; p = 0.157), indicating no significant difference in admission lactate elevation across polypharmacy groups.

### 3.2 Polypharmacy distribution and baseline characteristics

The median number of home medications was 11 (IQR 7–16). High polypharmacy (≥10 medications) was the most common category, affecting 1,970 patients (58.9%), followed by standard polypharmacy in 934 (27.9%) and no polypharmacy in 443 (13.2%). Patients with high polypharmacy were significantly older (median 71 vs 63 years), and had higher rates of hypertension (38.1% vs 28.2%), diabetes (24.2% vs 9.5%), chronic kidney disease (20.6% vs 7.4%), and congestive heart failure (24.3% vs 11.1%) compared with no polypharmacy. Importantly, SOFA scores did not differ significantly across polypharmacy groups (median 5 across all three; p = 0.875), confirming that polypharmacy captures a distinct dimension of risk beyond acute severity at ICU admission. Crude in-hospital ICU mortality was 18.5% (none), 26.0% (standard), and 27.5% (high polypharmacy; p < 0.001). Full baseline characteristics are presented in Table 1.

**Table 1.** Baseline characteristics by pre-admission polypharmacy category (N = 3,347)

| Variable | No polypharmacy<br>(0–4 meds) n = 443 | Standard polypharmacy<br>(5–9 meds) n = 934 | High polypharmacy<br>(≥10 meds) n = 1,970 | p-value |
| --- | --- | --- | --- | --- |
| **Demographics** |  |  |  |  |
| Age, years, median (IQR) | 63 (53–75) | 70 (59–82) | 71 (60–81) | <0.001 |
| Male sex, n (%) | 243 (54.9) | 513 (54.9) | 1,063 (54.0) | 0.865 |
| White race, n (%) | 290 (65.5) | 657 (70.3) | 1,439 (73.0) | 0.005 |
| **Comorbidities** |  |  |  |  |
| Hypertension | 125 (28.2) | 337 (36.1) | 750 (38.1) | <0.001 |
| Diabetes mellitus | 42 (9.5) | 157 (16.8) | 476 (24.2) | <0.001 |
| Chronic kidney disease | 33 (7.4) | 120 (12.8) | 406 (20.6) | <0.001 |
| Congestive heart failure | 49 (11.1) | 150 (16.1) | 478 (24.3) | <0.001 |
| COPD | 48 (10.8) | 95 (10.2) | 322 (16.3) | <0.001 |
| Charlson Comorbidity Index, median (IQR) | 0 (0–1.5) | 0 (0–2) | 0 (0–3) | <0.001 |
| **Severity at ICU admission** |  |  |  |  |
| SOFA score, median (IQR) | 5 (2–7) | 5 (2–7) | 5 (2–7) | 0.875 |
| Lactate (mmol/L), median (IQR) | 2.1 (1.9–2.6) | 2.1 (1.9–2.5) | 2.1 (1.8–2.4) | 0.005 |
| Lactate >2 mmol/L, n (%) | 320 (72.2) | 671 (71.8) | 1,357 (68.9) | 0.157 |
| Septic shock, n (%) | 165 (37.2) | 325 (34.8) | 649 (32.9) | 0.190 |
| Mechanical ventilation, n (%) | 439 (99.1) | 920 (98.5) | 1,953 (99.1) | 0.276 |
| **Infection source, n (%)** |  |  |  |  |
| Pulmonary | 76 (17.2) | 145 (15.5) | 358 (18.2) | 0.211 |
| Urinary tract | 37 (8.4) | 116 (12.4) | 241 (12.2) | 0.056 |
| Abdominal | 30 (6.8) | 62 (6.6) | 90 (4.6) | 0.030 |
| Bloodstream | 113 (25.5) | 222 (23.8) | 364 (18.5) | <0.001 |
| In-hospital ICU mortality, n (%) | 82 (18.5) | 243 (26.0) | 542 (27.5) | <0.001 |

IQR = interquartile range; SOFA = Sequential Organ Failure Assessment; COPD = chronic obstructive pulmonary disease. p-values: Kruskal-Wallis (continuous variables), chi-square (categorical variables). Mechanical ventilation rate of 99.0% reflects the near-universal intubation requirement in this tertiary ICU-admitted sepsis cohort.

### 3.3. Primary outcome: In-hospital ICU mortality

In multivariable logistic regression adjusting for age, sex, SOFA score, CCI, lactate, septic shock, mechanical ventilation, infection source, and time from ED to ICU, high polypharmacy was independently associated with in-hospital ICU mortality (aOR 1.45, 95% CI (1.10–1.91); p = 0.009) compared with no polypharmacy. Standard polypharmacy showed a borderline significant association (aOR 1.35, 95% CI (1.00–1.82); p = 0.051). Other independent predictors included SOFA score (aOR 1.21 per point, 95% CI (1.17–1.25)), CCI (aOR 1.12 per point, 95% CI (1.09–1.16)), lactate >2 mmol/L (aOR 1.64, 95% CI (1.31–2.04)), and urinary source (aOR 0.47, 95% CI (0.35–0.64)) and abdominal source (aOR 0.43, 95% CI (0.29–0.65)) compared with pulmonary sepsis. The full model AUROC was 0.722 compared with 0.656 for the SOFA-only model. Full regression results are presented in Table 2.

**Table 2.**
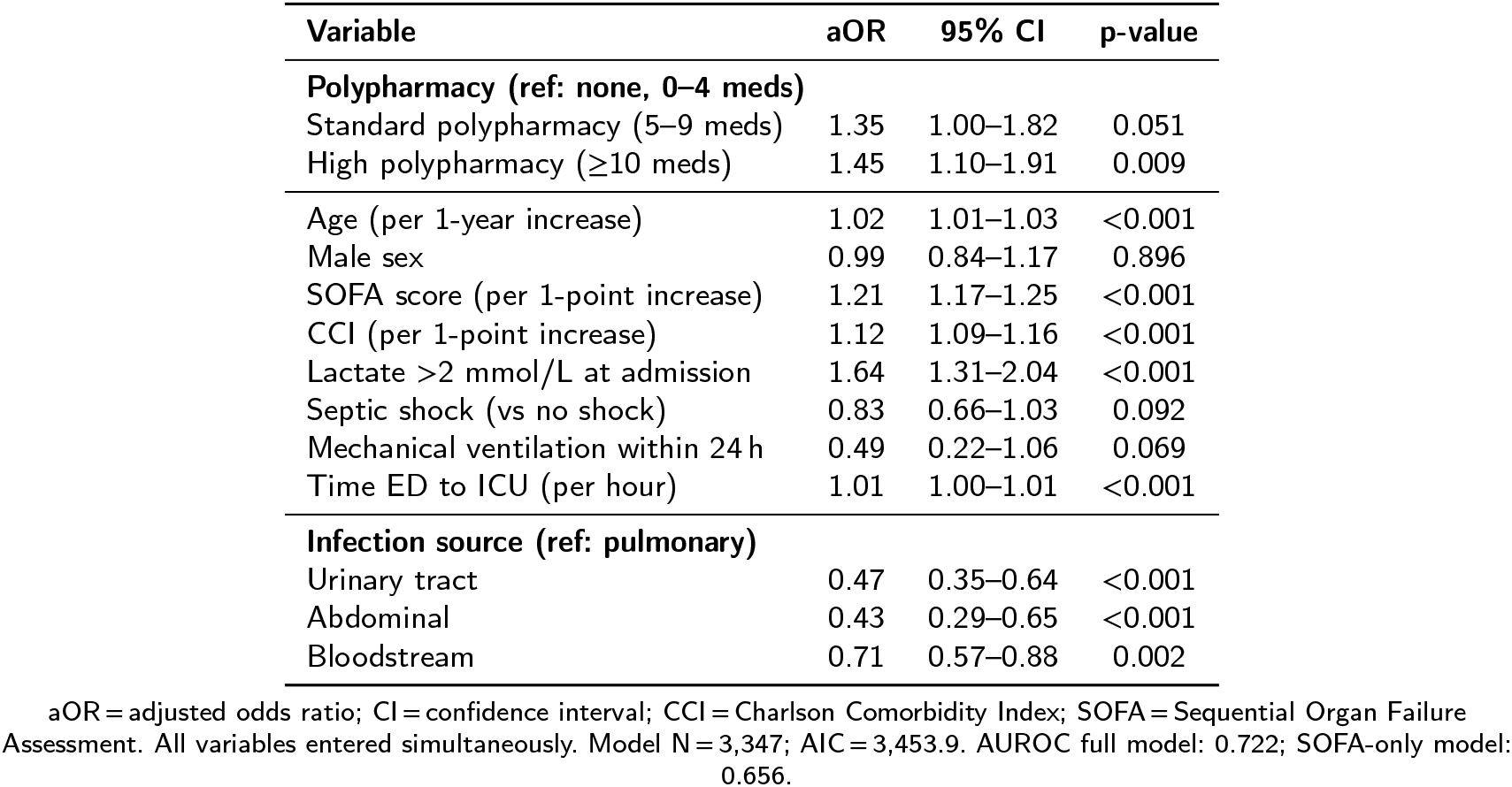
Multivariable logistic regression: independent predictors of in-hospital ICU mortality (N = 3,347)

| Variable | aOR | 95% CI | p-value |
| --- | --- | --- | --- |
| **Polypharmacy (ref: none, 0–4 meds)** |  |  |  |
| Standard polypharmacy (5–9 meds) | 1.35 | 1.00–1.82 | 0.051 |
| High polypharmacy ($\geq 10$ meds) | 1.45 | 1.10–1.91 | 0.009 |
| Age (per 1-year increase) | 1.02 | 1.01–1.03 | <0.001 |
| Male sex | 0.99 | 0.84–1.17 | 0.896 |
| SOFA score (per 1-point increase) | 1.21 | 1.17–1.25 | <0.001 |
| CCI (per 1-point increase) | 1.12 | 1.09–1.16 | <0.001 |
| Lactate >2 mmol/L at admission | 1.64 | 1.31–2.04 | <0.001 |
| Septic shock (vs no shock) | 0.83 | 0.66–1.03 | 0.092 |
| Mechanical ventilation within 24 h | 0.49 | 0.22–1.06 | 0.069 |
| Time ED to ICU (per hour) | 1.01 | 1.00–1.01 | <0.001 |
| **Infection source (ref: pulmonary)** |  |  |  |
| Urinary tract | 0.47 | 0.35–0.64 | <0.001 |
| Abdominal | 0.43 | 0.29–0.65 | <0.001 |
| Bloodstream | 0.71 | 0.57–0.88 | 0.002 |

aOR = adjusted odds ratio; CI = confidence interval; CCI = Charlson Comorbidity Index; SOFA = Sequential Organ Failure Assessment. All variables entered simultaneously. Model N = 3,347; AIC = 3,453.9. AUROC full model: 0.722; SOFA-only model: 0.656.

### 3.4 Dose-response analysis

Restricted cubic spline (RCS) regression demonstrated a significant, monotonically increasing relationship between the total number of home medications and the log-odds of in-hospital ICU mortality across the full range of observed values (Fig. 2). The relationship was significantly non-linear (likelihood ratio test p = 0.004), with a steeper increase in mortality risk observed above 8–9 medications. This dose-response pattern persisted after full adjustment for all prespecified covariates, held at their sample medians.

**Figure 2:**
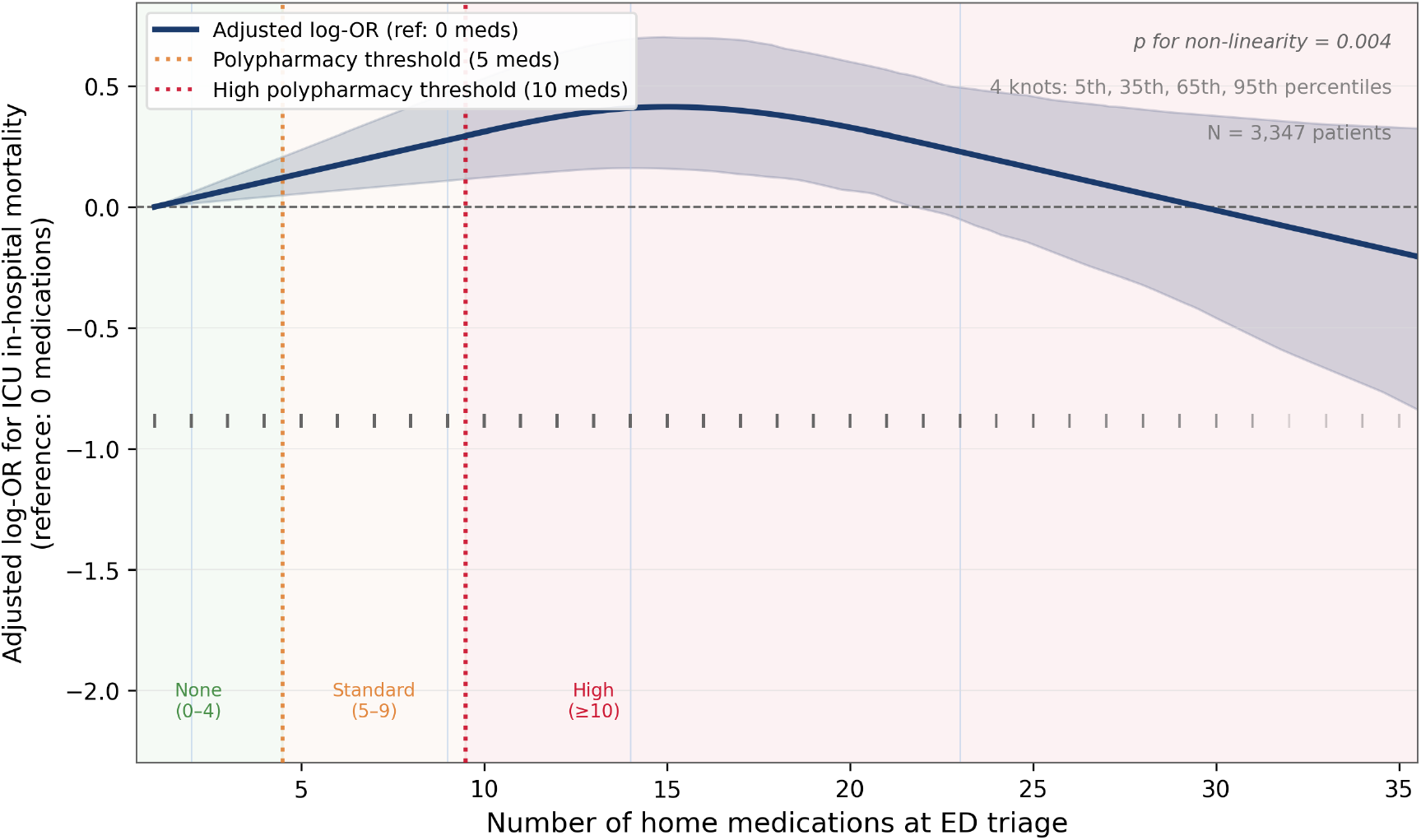
Restricted cubic spline analysis. Adjusted log-odds ratio for in-hospital ICU mortality as a function of continuous home medication count (N = 3,347). Spline fitted with four knots at the 5th, 35th, 65th, and 95th percentiles (knot positions: 2, 9, 14, 23 medications). Solid line = adjusted log-OR; shaded region = 95% bootstrap confidence interval (300 resamples). Reference at 0 medications; all covariates held at sample median. Orange dotted line = polypharmacy threshold (5 medications); red dotted line = high polypharmacy threshold (10 medications). Tick marks on the x-axis show the observed medication distribution (rug plot). The relationship is significantly non-linear (likelihood ratio test p = 0.004), with a steeper slope above 8–9 medications.

### 3.5 Secondary outcomes

High polypharmacy was significantly associated with 28-day mortality (aOR 1.47, 95% CI (1.14–1.91); p = 0.003) adjusted analysis (Table 3). By contrast, ICU LOS, hospital LOS, and new-onset AKI did not differ significantly by polypharmacy group in multivariable analysis. Despite a high AKI prevalence (50.6%), and near-universal mechanical ventilation (99.0%), no differences in LOS or AKI were observed. This likely reflects the overriding impact of illness severity in this highly selected critically ill cohort, whereby the independent effect of polypharmacy on resource utilization is diminished.

**Table 3.** Association between high polypharmacy (≥10 medications) and secondary outcomes, compared with no polypharmacy (0–4 medications)

| Outcome | Adjusted effect (95% CI) | Type | p-value |
| --- | --- | --- | --- |
| 28-day mortality | 1.47 (1.14–1.91) | aOR | 0.003 |
| ICU length of stay (days) | $\approx 0.0$ (−0.3 to 0.2) | Median diff. | 0.819 |
| Hospital length of stay (days) | $\approx 0.0$ (−0.9 to 1.0) | Median diff. | 0.922 |
| New-onset AKI (KDIGO) | 1.10 (0.88–1.38) | aOR | 0.398 |

aOR = adjusted odds ratio; AKI = acute kidney injury (KDIGO criteria). All models adjusted for age, sex, SOFA score, CCI, lactate, septic shock, and mechanical ventilation. LOS differences estimated via quantile regression at the median. Overall ICU LOS: median 2.8 days (IQR 1.8–5.3); hospital LOS: median 9.8 days (IQR 5.8–17.6); overall AKI incidence: 50.6%.

### 3.6 Drug class sub-analysis

Within the high polypharmacy group (n = 1,970), drug-class analysis revealed several striking patterns (Table 4). Statins were present in 57.5% of patients and were independently associated with markedly reduced mortality risk (aOR 0.56, 95% CI (0.45–0.70); p < 0.001), consistent with pleiotropic anti-inflammatory effects. Antidiabetic agents (aOR 0.66, 95% CI (0.53–0.83)) and beta-blockers (aOR 0.75, 95% CI (0.61–0.93)) were also associated with lower mortality. Conversely, RAS agents were associated with reduced mortality when analysed alone (aOR 0.67, 95% CI (0.54–0.82)), but the combination of RAS blockers plus diuretics (present in 30.1% of patients) was associated with significantly increased AKI risk (aOR 1.49, 95% CI (1.21–1.84); p < 0.001). These findings suggest that while many drug classes individually carry protective associations, their concurrent combination in the polypharmacy stratum drives the net adverse outcome effect through complex pharmacodynamic interactions.

**Table 4.** Drug class sub-analysis within the high polypharmacy group (n = 1,970): association of home medication classes with ICU in-hospital mortality, adjusted for SOFA score, age, sex, CCI, septic shock, and mechanical ventilation.

| Drug class | Present n (%) | Absent n (%) | aOR (95% CI) | p-value |
| --- | --- | --- | --- | --- |
| RAS agents (ACE inhibitor / ARB) | 897 (45.5) | 1,073 (54.5) | 0.67 (0.54–0.82) | <0.001 |
| Diuretics | 1,093 (55.5) | 877 (44.5) | 0.82 (0.66–1.01) | 0.058 |
| Anticoagulants | 665 (33.8) | 1,305 (66.2) | 0.72 (0.57–0.90) | 0.004 |
| Systemic corticosteroids | 670 (34.0) | 1,300 (66.0) | 0.89 (0.71–1.11) | 0.308 |
| Immunosuppressants | 174 (8.8) | 1,796 (91.2) | 1.13 (0.78–1.63) | 0.521 |
| Antidiabetic agents | 750 (38.1) | 1,220 (61.9) | 0.66 (0.53–0.83) | <0.001 |
| Beta-blockers | 1,219 (61.9) | 751 (38.1) | 0.75 (0.61–0.93) | 0.010 |
| Statins | 1,133 (57.5) | 837 (42.5) | 0.56 (0.45–0.70) | <0.001 |
| RAS blockers + diuretics<sup>†</sup> | 592 (30.1) | 1,378 (69.9) | 1.49 (1.21–1.84) | <0.001 |

aOR = adjusted odds ratio; CI = confidence interval; RAS = renin-angiotensin system; ACE = angiotensin-converting enzyme; ARB = angiotensin receptor blocker.
<sup>†</sup>Last row reports aOR for *new-onset AKI* (secondary outcome), not mortality. Percentages refer to proportions within the high polypharmacy group (n = 1,970).

### 3.7 Propensity score-matched analysis

Of 1,970 high polypharmacy patients, 1,377 control patients (none/standard polypharmacy) were available for matching. After 1:1 nearest-neighbour propensity score matching with replacement, 1,969 matched pairs were formed within the caliper (1,377 unique controls, some matched to more than one treated patient). All covariates were well-balanced post-matching (maximum SMD = 0.074, all below the 0.10 threshold). In the matched cohort, in-hospital ICU mortality was 27.5% (high polypharmacy) versus 23.1% (controls). Logistic regression confirmed the primary finding (matched aOR 1.28, 95% CI (1.10–1.50); p = 0.001), supporting robustness of the association to residual confounding by indication.

### 3.8 Subgroup analyses

Subgroup analyses by age, sex, septic shock status, mechanical ventilation, and infection source demonstrated consistent point estimates across all prespecified subgroups, with aORs ranging from 1.01 to 1.30 (Fig. 3). The association was statistically significant in the overall cohort (aOR 1.20, 95% CI (1.01–1.41); p = 0.035), in the noseptic-shock subgroup (aOR 1.30, 95% CI (1.04–1.61); p = 0.018), among mechanically ventilated patients (aOR 1.20, 95% CI (1.02–1.42); p = 0.031), and in non-pulmonary infection sources (aOR 1.24, 95% CI (1.03–1.49); p = 0.025). The septic shock subgroup could not be computed due to near-complete separation (collinearity between septic shock and vasopressor prescriptions producing a singular design matrix). No significant interaction term was detected across any subgroup (all p > 0.10 after Bonferroni correction).

**Figure 3:**
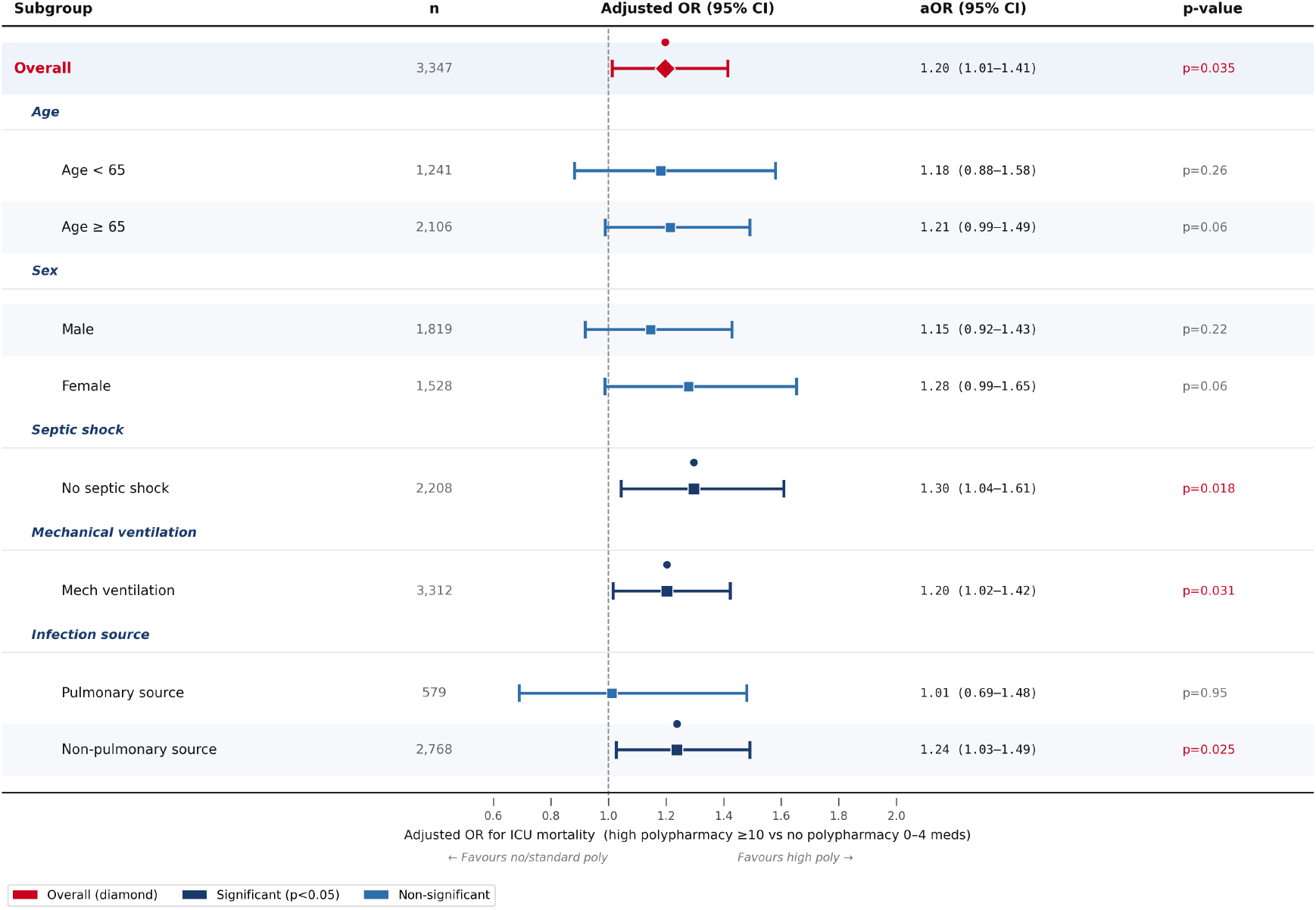
Subgroup forest plot. Adjusted ORs for in-hospital ICU mortality associated with high polypharmacy (≥10 medications) versus no polypharmacy (0–4 medications) across prespecified subgroups. Each estimate derives from a logistic regression model adjusted for age, SOFA score, CCI, and lactate. The overall estimate presented here (aOR = 1.20; 95% CI (1.01–1.41); p = 0.035) represents a partially adjusted model to maintain consistency with the subgroup models, whereas the primary fully adjusted model (Table 2) yielded an aOR of 1.45. The septic shock subgroup was omitted due to model non-convergence (near-complete separation with vasopressor data). Filled diamond = overall estimate; filled squares = subgroup estimates; navy = p < 0.05; medium blue = p ≥ 0.05. Subgroup sample sizes: Overall n = 3,347; Age <65 n = 1,241; Age ≥65 n = 2,106; Male n = 1,819; Female n = 1,528; No septic shock n = 2,208; Mech ventilation n = 3,312; Pulmonary source n = 579; Non-pulmonary source n = 2,768.

### 3.9 Incremental predictive value

Addition of polypharmacy classification to a SOFA-only model improved the AUROC from 0.656 to 0.662, with a continuous NRI of 0.24 and IDI of 0.005. Adding polypharmacy to the SOFA+CCI model further improved AUROC from 0.669 to 0.673 (NRI = 0.14; IDI = 0.004). These improvements, while modest in absolute magnitude, were consistent across both base models and suggest that medication burden provides incremental prognostic information beyond established severity scores-information available immediately at ED presentation.

## 4. Discussion

In this retrospective cohort study of 3,347 ICU patients with sepsis, we demonstrate that pre-admission high ypharmacy (≥10 medications) is independently associated with a 45% increase in the adjusted odds of in-hospital ICU mortality (aOR 1.45, 95% CI (1.10–1.91); p = 0.009) and a 47% increase in 28-day mortality (aOR 1.47, 95% CI (1.14–1.91); p = 0.003), after comprehensive adjustment for SOFA score, Charlson Comorbidity Index, septic shock, and mechanical ventilation. These findings were confirmed in propensity score-matched analysis (matched aOR 1.28; p = 0.001) and were consistent across prespecified subgroups. To our knowledge, this is the first study to use the medrecon table in MIMIC-IV-ED, capturing real-world home medication lists at ED triage, to establish polypharmacy as an independent predictor of ICU mortality in sepsis.

The finding that SOFA scores did not differ across polypharmacy groups (median 5 in all three; p = 0.875) is central to the interpretation of our results. It confirms that polypharmacy captures a dimension of risk that is distinct from acute physiological derangement at ICU admission. This is consistent with the hypothesis that polypharmacy serves as a proxy for long-term physiologic vulnerability, what some authors have termed the ‘pharmacologic frailty’ phenotype, that predisposes patients to worse sepsis outcomes through mechanisms that precede and persist beyond the acute presentation [14, 15].

The drug class analysis provided mechanistically informative findings. The strong protective association of statins (aOR 0.56; p < 0.001) is consistent with extensive prior literature on the pleiotropic anti-inflammatory effects of HMG-CoA reductase inhibitors in critical illness, including modulation of endothelial activation, neutrophil function, and cytokine release [16]. The protective signals for antidiabetic agents (aOR 0.66) and beta-blockers (aOR 0.75) may reflect that patients prescribed these agents have well-managed metabolic and cardiovascular disease, representing a relatively ‘controlled’ polypharmacy phenotype with preserved physiologic reserve. Notably, systemic corticosteroids showed no significant mortality association (aOR 0.89; p = 0.308), though home corticosteroid use has been linked to critical illness-related corticosteroid insufficiency [17]. Conversely, the combination of RAS blockers plus diuretics significantly increased AKI risk (aOR 1.49), consistent with their known interaction to impair renal autoregulation under systemic hypoperfusion during sepsis [6, 18]. This finding has direct clinical relevance: patients presenting to the ED with this combination on their medication list may benefit from urgent pharmacy review and temporary discontinuation at ICU admission.

The incremental predictive value of polypharmacy beyond SOFA score, while modest (NRI = 0.24; ΔAUROC=+0.006), is clinically meaningful in its accessibility. Unlike SOFA, which requires laboratory values and clinical assessment that may take hours to complete after ICU admission, the medication count from the ED triage reconciliation is available immediately upon patient presentation. In time-critical sepsis management, this makes polypharmacy count a unique early risk-stratification signal.

The absence of a significant effect on ICU LOS and AKI in adjusted analysis, despite a significant mortality signal, may partly reflect the overwhelmingly high prevalence of mechanical ventilation (99.0%) in this cohort. When virtually patients require intubation, determinants of LOS become dominated by ventilation duration and weaning success rather than pre-admission medication burden. Similarly, with AKI present in 50.6% of patients overall, the already-high baseline AKI risk in this ICU sepsis population may attenuate detectable additional contributions from polypharmacy.

Our study has several limitations. First, as a single-centre dataset from a US tertiary academic centre, findings may not generalise to community hospitals or low-resource settings. Second, medication reconciliation completeness may vary for patients who were intubated or unconscious at triage. Third, polypharmacy count reflects documented or prescribed medications rather than confirmed adherence. Fourth, the sepsis cohort was defined using ICD codes rather than the full computational Sepsis-3 algorithm, which may introduce misclassification. Fifth, residual confounding by unmeasured frailty and functional status cannot be excluded. Sixth, the near-universal mechanical ventilation (99.0%) in this cohort, substantially higher than most ICU populations, likely reflects BIDMC’s tertiary referral case mix and limits generalisability to less severe sepsis populations.

## 5. Conclusion

This study demonstrates that pre-admission high polypharmacy is an independent predictor of ICU and 28-mortality in sepsis, providing prognostic information beyond established severity indices such as SOFA and comorbidity burden. Leveraging the underutilised medication reconciliation data from the MIMIC-IV-ED database, this work is, to our knowledge, the first to quantify the independent contribution of medication burden to critical care outcomes in sepsis using real-world triage-level data. Importantly, medication count is available immediately at emergency department presentation, making it a practical and scalable tool for early risk stratification before full clinical and laboratory assessment is complete. The identification of specific high-risk medication patterns, such as the combination of RAS blockers and diuretics, further highlights actionable opportunities for targeted intervention. These findings support the integration of structured medication review and pharmacist-led optimisation into early ICU care pathways. Overall, this study positions polypharmacy not merely as a marker of multimorbidity, but as a measurable and potentially modifiable determinant of outcomes in sepsis. Incorporating medication burden into clinical decision support systems and prospective interventional strategies may enhance early risk prediction and improve patient outcomes in this high-risk population.

## Ethical approval

This study uses the MIMIC-IV and MIMIC-IV-ED databases, which are de-identified and publicly available via PhysioNet under an approved data use agreement. No new human participants were recruited and no animals were involved. Institutional review board approval was not required.

## Data Availability Statement

MIMIC-IV (v2.2) and MIMIC-IV-ED (v2.2) are freely available at https://physionet.org upon completion of a credentialling application and CITI training.

